# COVID-19 deaths detected in a systematic post-mortem surveillance study in Africa

**DOI:** 10.1101/2020.12.22.20248327

**Authors:** Lawrence Mwananyanda, Christopher J. Gill, William MacLeod, Geoffrey Kwenda, Rachel Pieciak, Zachariah Mupila, Francis Mupeta, Leah Forman, Luunga Ziko, Lauren Etter, Donald Thea

## Abstract

**Objectives:** Limited SARS CoV 2 testing in many African countries has constrained availability of data on the impact of COVID-19 (CV19). To address this gap, we conducted a systematic post-mortem surveillance study to directly measure the fatal impact of CV19 in an urban African population.

**Design:** We enrolled deceased individuals at the University Teaching Hospital (UTH) Morgue in Lusaka, Zambia. We obtained nasopharyngeal swabs for testing via reverse-transcriptase quantitative PCR (RT-qPCR) against the SARS-2 Coronavirus. We stratified deaths by CV19 status, by location, age, sex, and underlying risk factors.

**Setting:** UTH is Zambia’s largest tertiary care referral hospital and its morgue registers ∼80% of Lusaka’s deaths.

**Participants:** Participants of all ages were enrolled if within 48 hours of death and if the next of kin or representative provided written informed consent.

**Results:** We enrolled 372 participants between June and September 2020, and had PCR results for 364 (99.5%). CV19 was detected in 70/364 (19.2%). The median age for CV19+ deaths was 48 years (IQR 36-72 years) and 70% were male. Most CV19+ deaths (51/70, 72.8%) occurred in the community; none had been tested for CV19 antemortem. Among the 19/70 facility deaths, six were tested antemortem. Among the 52/70 CV19 deaths with symptoms data, 44/52 had typical symptoms of CV19 (cough, fever, shortness of breath), of whom only five were tested antemortem. We identified CV19 among seven children; only one had been tested antemortem. The proportion of CV19+ deaths increased with age, but 75.7% of CV19+ deaths were aged <60 years. The five most common co-morbidities among CV19+ deaths were: tuberculosis (31.4%); hypertension (27.1%); HIV/AIDS (22.9%); alcohol use (17.1%); and diabetes (12.9%).

**Conclusions:** Contrary to expectations, CV19+ deaths were common in Lusaka. The majority occurred in the community where testing capacity is lacking. Yet few who died at facilities were tested, despite presenting with typical symptoms of CV19. Therefore, CV19 cases were under reported because testing was rarely done, not because CV19 was rare. If our data are generalizable, the impact of CV19 in Africa has been vastly underestimated.

## INTRODUCTION

The global COVID-19 (CV19) pandemic has had a devastating impact on the health of millions, with ongoing and/or accelerated transmission in nearly all regions of the world.^1^ A puzzling question is why CV19 appears to have largely skipped Africa?^2-4^ CV19 transmission should be favored wherever population densities are high, yet the institute of Health Metrics and Evaluation (IHME) recently listed CV19 as the 45^th^ most common cause of death in Africa, despite being the 12^th^ leading cause globally.^5^ There is no obvious reason why CV19 would not spread as efficiently in Nairobi, Accra, or Lagos as it has in New York City, London, or Mumbai. Currently, most of the systematic evidence about CV19’s impact in Africa comes from South Africa, which has documented >750,000 cases of CV19, >20,000 deaths and a case fatality rate of 2.7%.^6-8^

The so-called ‘Africa paradox’ has been the focus of a number of editorials in prominent journals^9-13^ proposing several explanatory theories. These include: 1) exposure to non-CV19 coronaviruses has induced cross-reactive herd immunity;^14^ 2) the younger age structure of African populations allows its populations to better tolerate CV19;^15^ 3) experience gained during the Ebola crisis allowed public health agencies across Africa to better contain CV19;^16^ and 4) that certain live attenuated vaccines - Bacille Calmette Guerin (BCG) vaccine, the oral polio vaccine and measles vaccines - engendered robust non-specific innate immune responses that also protect against CV19.^17-19^

We postulate a more mundane explanation for the Africa paradox: insufficient data. Africa is the world’s poorest continent. Disease surveillance is resource intense and takes time to establish *de novo*, challenges that are magnified in a global health emergency. CV19 deaths might be challenging to identify among countries with limited resources to establish new disease surveillance systems, particularly in countries with already high background mortality rates.

Since 2017, our team has been conducting systematic post-mortem surveillance for respiratory pathogens among deceased infants in Lusaka, Zambia. With the onset of the CV19 pandemic, we were able to quickly amend our study to expand surveillance to include all age groups and to test for CV19. We present results from the first three months of surveillance. Counter to expectations, CV19 deaths proved to be common.

## METHODS

### Study setting

According to the World Bank, Zambia is a poor country that ranked 117^th^ out of 128 countries in terms of economic competitiveness in 2007.^20^ Its economy has been stalled since 2015 due to falling copper prices, and with the CV19 pandemic, its economy was predicted to contract in 2021 by 4.5%.^20^ Zambia’s public debt burden is 60% of its gross domestic product, and over 50% of Zambians live below the poverty line.^21^ The capital Lusaka has ∼2 million citizens and is Zambia’s largest city.^22^ In Lusaka, the majority of the urban poor live in densely crowded peri-urban slums where there are few places to avoid crowds, making it difficult to practice social spacing. According to the study staff, masks were difficult to obtain and seldom worn by the general Lusaka population during this time.

### Overview of the ZPRIME study and its expansion to CV19 surveillance

The Zambia Pertussis RSV Infant Mortality Estimation study (ZPRIME) was launched in August 2017 to measure the post-mortem prevalence of these two diseases among young infants in southern Africa (Lusaka, Zambia). While Lusaka has dozens of health facilities, few can issue the death certificates that are legally required to inter a body. For the CV19 expansion, we concentrated our resources at the University Teaching Hospital (UTH) morgue. UTH is the primary tertiary care referral hospital in Lusaka and registers at least 80% of deaths in the city, including those from UTH itself and from the community.

In June 2020, we amended our protocol to expand enrollment to deceased individuals of all ages and to add CV19 PCR testing. Other than these changes of scope, ZPRIME’s protocol did not change. Ethical oversight for ZPRIME and the CV19 expansion were provided by the Institutional Review Boards at Boston University and the University of Zambia. Written informed consent was obtained from the deceased’s family members or representatives.

For the CV19 study expansion, we enrolled deceased individuals who either died in the community or at a facility in Lusaka. We define ‘facility deaths’ as those that occurred under care at UTH. All other deaths were defined as ‘community deaths’, which may have included deaths referred to UTH from smaller facilities in Lusaka. From each deceased individual, we tabulated sex, age, location of death (facility vs. community), geographic location of the death within Lusaka, presenting symptoms, underlying risk factors, and the results of antemortem CV19 PCR testing (if any).

### Enrollment procedures

Enrollment occurred as next of kin came to claim the bodies of their loved ones at the UTH morgue. Our only exclusion criterion was enrollment > 48 hours after death to reduce the risk of false negative PCR results from degradation of viral RNA. An abbreviated grief counselling session was offered unconditionally to the family members. Due to the high volume of deaths, and our team’s finite capacity, we only enrolled Monday through Friday, 9AM to 5PM. During these times, we enrolled every fifth death during July and every third death in August, with a daily cap of ∼5 deaths per day in both cases. In September, after the ZPRIME infant study ended allowing our full team to focus on the CV19 project, we enrolled at a 1:1 ratio without a daily cap. Since the enrollments were done prior to assessing clinical data, such as symptoms at presentation, and prior to CV19 testing, this approach should not have introduced any selection bias.

Presenting respiratory symptoms were solicited from informants from both facility and community deaths. For facility deaths, additional clinical data were extracted from the medical chart and from the official death certificate. For deaths occurring in the community, we used an abbreviated verbal autopsy tool to identify respiratory symptoms and underlying medical conditions.^23^ This included a free text narrative that allowed the next of kin to describe the circumstances of each death. In this way, we attempted to characterize the clinical syndrome preceding the death and to identify underlying risk factor co-morbidities that might be associated with CV19 mortality. This included previously described risk factors, such as hypertension, heart disease, and diabetes, but also putative risk factors prevalent in Africa including HIV/AIDS, tuberculosis, malnutrition and sickle cell disease. Other conditions, such as epilepsy and malaria, were added iteratively as we reviewed the clinical data from each of the deceased.

### Sample collection

Nasopharyngeal (NP) samples were taken using flocked-tipped nylon swabs (Copan Diagnostics, Murrieta CA), sized for infants or adults as required.^24 25^ Swabs were inserted into both nares, advanced until they reached the posterior nasopharynx and then rotated 180 degrees in both directions. Samples were immediately placed in universal transport media on ice or in cool boxes at 4-8C, and transported to our on-site PCR lab for accession, aliquoting and storage at −80C. Our lab is located ∼50 yards from the morgue, allowing samples to be transported within minutes of collection.

### Laboratory procedures

After vortexing to remove sample from the NP swabs, total nucleic acid (TNA) was extracted using the NucliSens EASYMAG system (bioMerieux, Marcy l’Etoile, France).^26^ We used reverse transcriptase quantitative PCR (RT-qPCR) to identify the SARS2 corona virus that causes CV19. Due to difficulties and delays in obtaining PCR kits in Zambia, the first 16 PCR runs used the US CDC kit that target the N1 and N2 regions of the virus’ nucleocapsid protein gene. Runs 17-18 used a different kit that targeted a consensus sequence of the nucleocapsid gene [NCBI Reference Sequence: NC_045512.2] and the gene sequence encoding an overlapping polyprotein that is later cleaved *in vivo* into PP1ab and PP1a polyproteins [NCBI Gene ID: 43740578] (Wuhan EasyDiagnosis Biomedicine Co, Ltd, Wuhan, China).

For both assays, a positive reaction against the respective gene targets was defined as a cycle threshold (CT) <40. PCR was run for 45 cycles, making a CT of 45 the effective limit of detection. We also included all individuals with a CT<45 in this analysis. Given concerns of higher false positivity results using the latter cutpoint, we report our results at each threshold.

In addition to running positive and negative controls on each assay plate, both assays included qPCR against the constitutively expressed human RNAseP gene for each sample as a quality control measure demonstrating adequacy of sample collection and nucleic acid extraction, and the absence of PCR inhibition.

### Analytic approach

We calculated a simple prevalence of CV19 by dividing the number of deaths where CV19 was detected by all deaths in the enrolled sample. We conducted no modeling and did not use imputation. We stratified deaths by sex, age, geography, and location (facility vs. community deaths). A key distinction is that individuals who died at UTH could have been tested for CV19 antemortem, whereas there was far more limited capacity for CV19 testing in the community.

To examine the distribution of age at death, we cataloged age-tratified death statistics from the Zambian government’s official burial registry during the surveillance period. This allowed us to construct a contemporaneous death-by-age distribution to infer whether our enrolled sample was representative of all deaths during this period and whether the subset of CV19+ deaths followed a similar distribution.

We used ArcGIS (Esri Inc, Redlands, CA) software to map the locations of the deaths within Lusaka. We pulled population size data from the Zambia Data Hub: https://zambia-open-data-nsdi-mlnr.hub.arcgis.com/, which is managed by the Government of Zambia through the Ministry of Lands and Natural Resources, and the Zambia Statistics Agency. We downloaded these ArcGIS layers to the level of Lusaka’s city wards.

Inferring causality is a challenge in post mortem studies: CV19 may have a direct or indirect role in deaths, or could be coincidental. The US CDC has issued guidance stating that in deaths where CV19 is detected, the virus should be assumed to be the direct or underlying cause of death, absent exonerating circumstances.^27^ To provide additional insight into causality, we sorted CV19+ deaths into four categories based on clinical presentation data reported contemporaneously in the medical chart and/or death certificate (facility deaths) or from the verbal autopsy (community deaths):

1. <u>Probable CV19</u>, which included those with any combination of witnessed or reported cough, fever, upper respiratory symptoms, shortness of breath, or difficulty breathing.
2. <u>Possible CV19</u>, which included individuals presenting with symptoms that may reflect common sequellae of CV19, such as apparent or confirmed stroke, myocardial infarction, sudden onsent abdominal pain, nausea, vomiting or diarrhea;
3. <u>Probably not CV19</u>, which included patients for whom medical data indicated a non-CV19 cause of death; and
4. <u>Uncertain</u>, for individuals lacking sufficient data to allow for adjudication.

## RESULTS

### Summary of testing results

Between June 15 and Oct 01, 2020, our team enrolled 372 deceased individuals, ranging in age from <1 year to 105 years. Consent rates were 99.5%, and the main reason for non-consent was among Muslim families who cited conflicts with their cultural burial practices. Ten samples could not be matched to enrollments, leaving 362/372 (97.3%) with samples for testing. In addition, we included two individuals who had been tested positive for CV19 antemortem outside of the study, for a total of 364 deaths in our analytic sample. For the 362 tested in our lab, all had RNAseP results with CT<35 indicating adequacy of the sampling collection and DNA extraction processes. We did not have RNaseP results for the two individuals tested antemortem at a different lab.

Of these 364 deaths, 96 (25.8%) were facility deaths and 268 (74.2%) were from the community. Contemporaneously, there were 3,676 deaths recorded in the burial registry between June 15 and September 30, 2020, meaning that the enrolled sample represented ∼10% of total deaths at the UTH morgue. Consistent with the distribution of deaths in the enrolled sample, the majority of CV19+ were community deaths (51/70, 72.8%), while only 19/70 (28.6%) were facility deaths.

Using the stricter PCR threshold of a CT<40, 58/364 (15.9%) were CV19 positive. When expanding that to individuals with any detectable level of the virus (i.e., CT values from 40 to 45), we identified a further twelve CV19+ deaths, bringing the total to a presumptive prevalence of CV19 in 70/364 cases (19.2%).

Among the 70 CV19+ deaths, antemortem testing for CV19 had rarely been performed. Among the facility deaths, only 6/19 (31.6%) had been tested antemortem. Among the majority of 51/70 that were community deaths, none had been tested for CV19 antemortem.

### Distribution of CV19 deaths by time, sex, age, and geography

CV19+ deaths were detected throughout the entire period of observation, with notable week to week variation over time (**Figure 1**). As a proportion of deaths, CV19 deaths were most common in late July to early August 2020.

**Figure 1.**
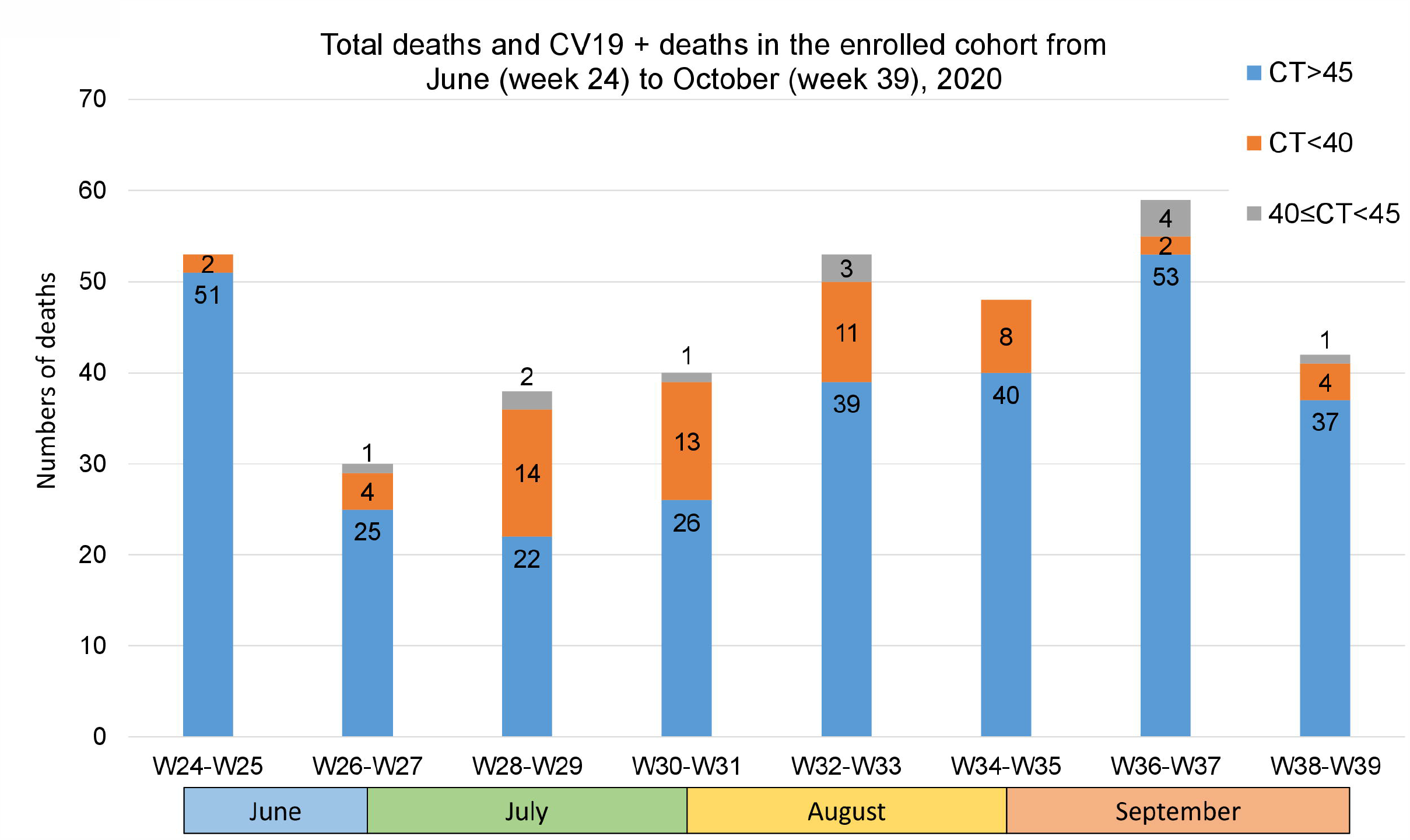
Bi-Weekly detection of COVID-19 deaths, June – September 2020. CV19+ cases are presented at two cycle thresholds: <40 (in orange) and the additional cases detected at a CT value between 40 and 45 (in grey), against total deaths (in blue). For simplicity, we present these with cases clustered in 2 week increments, from calendar weeks 24 through 39. As can be seen, detection of COVID-19 occurred throughout the period of surveillance, though with significant week to week variability. Due to the high volume of deaths at the morgue, and the ongoing needs to complete our work on the ongoing post mortem infant study that continued through August 31, 2020, we were only able to enroll a subset of deaths on a daily basis. Therefore, in July we enrolled every fifth individual, capping enrollments at around 5-6 participants per day; in August we reduced to enroll every third invididual, with the same daily cap; and in September, when our full team could focus on the CV19 cases, we expanded to a 1:1 enrollment. For this reason, the total number of enrolled deaths in July and August represent only about a tenth of deaths that occurred during those periods. This has no bearing on our prevalence calculations, but does mean that the absolute number of deaths will be undercounted in this figure.

Overall, 70% of the CV19+ deceased were male, with a similar male to female ratio among the facility and community deaths (**Figure 2**). By comparison, in the burial registry, 60% of deaths were male and 40% were female. Thus men appeared to be over-represented among CV19+ deaths.

**Figure 2.**
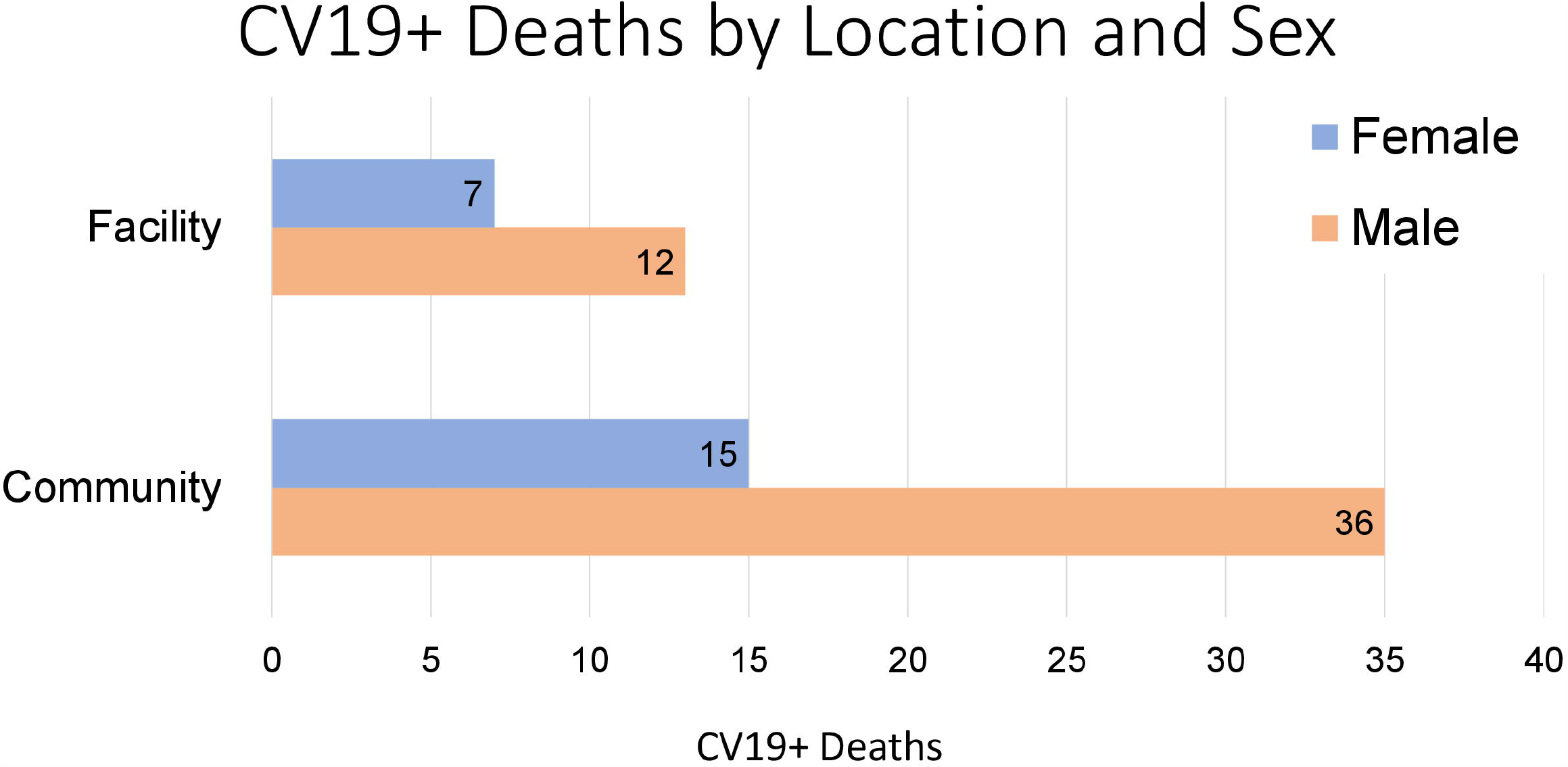
Distribution of COVID-19 deaths by sex and facility vs. community setting. COVID-19 was detected far more often among males than females. 74% of all deaths occurred in the community, and therefore were systematically excluded from antemortem CV19 testing. This is obviously a significant factor in explaining why CV19 is being undercounted. Yet, CV19 testing among the facility deaths was also uncommon, despite the fact that most participants had presented with typical symptoms suggestive of CV19, such as cough, fever, and difficulty breathing.

The median age among the CV19 deaths was 48 years (IQR 36-72 years). The median age among community deaths was 47 years (IQR 34-72 years), and was somewhat older among facility deaths at 55 years (IQR 38-73 years). **Figure 3a** provides the numbers of deaths across the age distribution, while **Figure 3b** presents these as a proportion of deaths that were CV19+ within each stratum. There were several deaths in children < 19 years, within which we observed 5 (or 7) CV19+ deaths, depending on the CT threshold.

**Figure 3.**
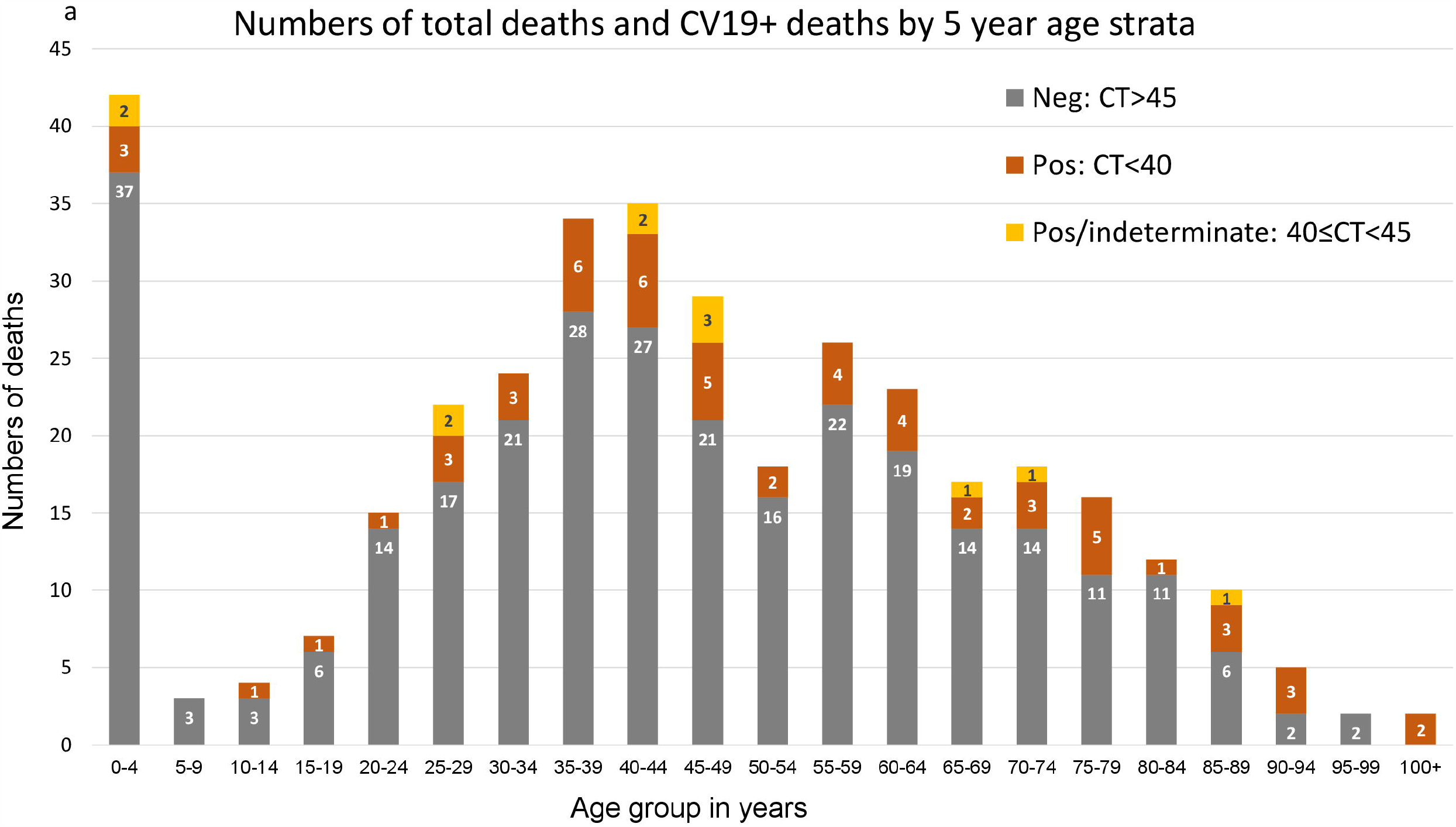
Distribution of CV19 deaths by five year age increments. **Figure 3a Numbers of deaths by age group** presents the numbers of deaths by PCR testing result. In blue were deaths without CV19; in orange are those where CV19 was detected at a CT value <40; in grey are the additional cases where CV19 was detected at a CT between 40 and 45. The results falling in the 40-45 range would be considered ‘indeterminate’ results, but that does not mean that they are false positives. Given the high index of suspicion, a more parsimonious explanation is that they represent true positives, albeit at a lower signal intensity. This could reflect natural biological variation (e.g, waning signal intensity at the end of the arc of an infection) or variations in sample collection, sample degradation over time, or laboratory processes.

**Figure 3b.**
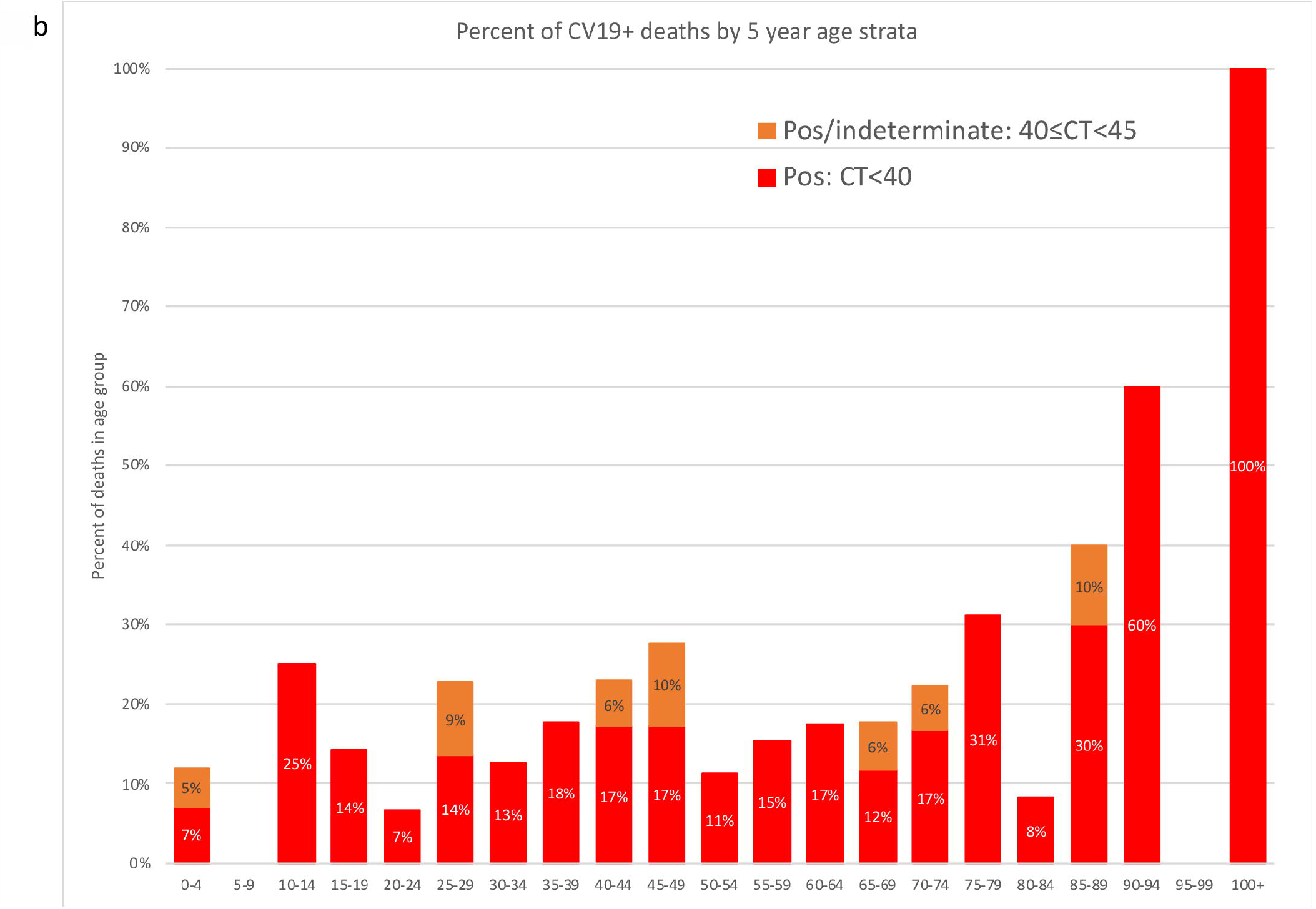
Proportions of deaths by age group. presents these same data but as proportions of positive results, at both thresholds, by age strata. We note that the highest proportions of CV19 deaths / total deaths are seen clustered in older persons. However, the highest number of cases is concentrated among those <60 years, with similar proportions of CV19 detections across most age strata.

While the proportion of CV19 deaths out of total deaths increased above the age of 70 years, in absolute numbers, the majority (53/70, 75.7%) of all CV19 deaths were in individuals <60 years, and clustered between 20 and 59 years (46/70, 65.7%). To explore this further, we compared the distribution of deaths by age among the deaths in our sample, the 70 among whom CV19 was detected in our lab, and the complete set of >3500 deaths that were registered at the burial office during the study period (**Figure 4**). Overall, the age distribution for the enrolled cohort closely matched the age distribution from the complete set of deaths from the burial registry, suggesting that our sample was representative of the overall distribution of deaths by age. By contrast, the CV19+ deaths were somewhat underrepresented among those aged 0-39 years, and somewhat overrepresented among those aged 60 and above.

**Figure 4.**
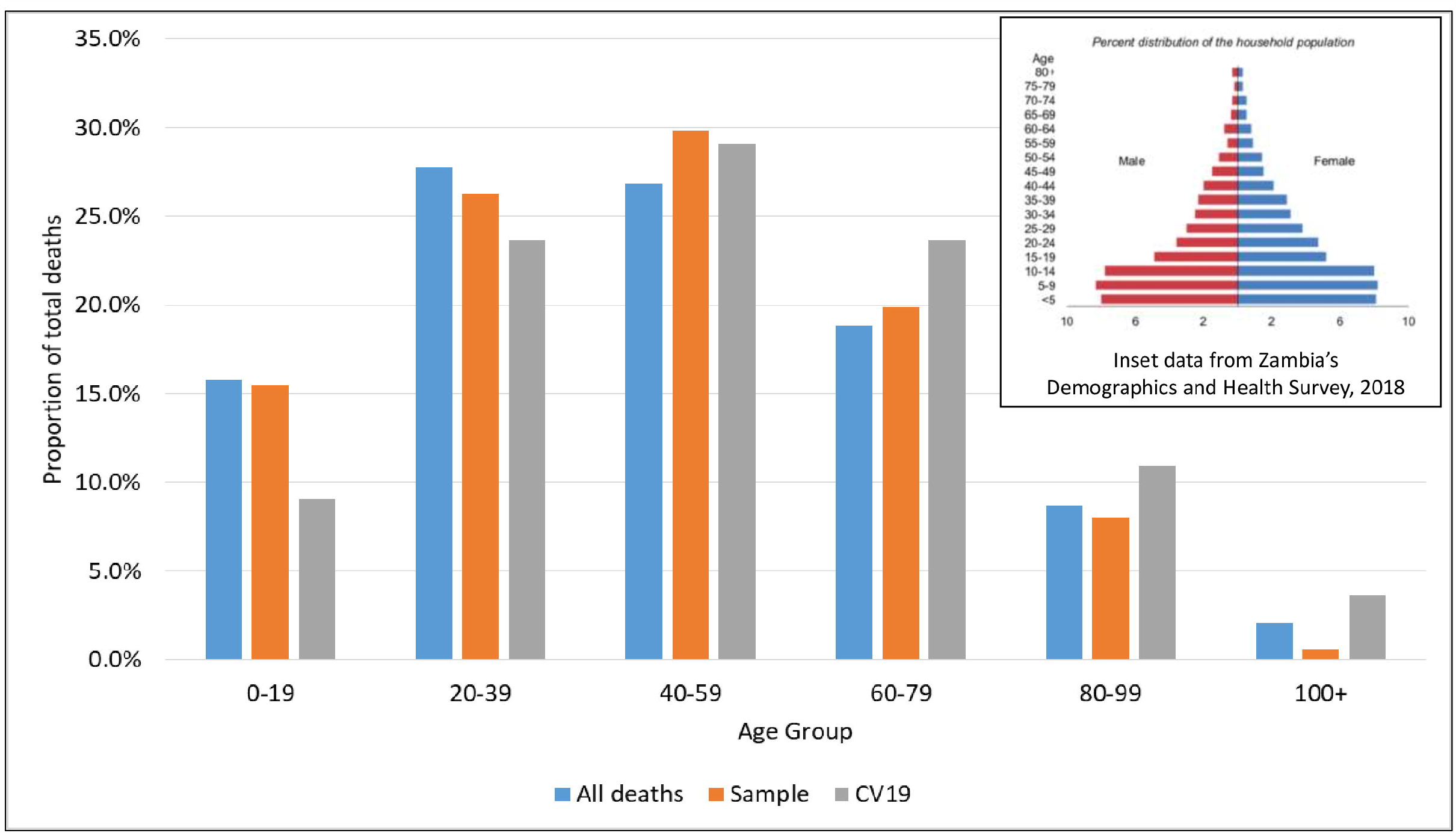
Comparison of age distribution among total enrolled deaths vs. registered deaths vs. CV19+ deaths This figure shows the age distribution of the three populations assessed in this analysis, in 5-year age strata. In blue are all of the deaths in the enrolled sample; in orange are the ages from the burial registry, representing all deaths that occurred during the sampling period; in grey are the deaths within the CV19+ sub-sample. For reference, in the inset we provide the population age structure, by sex, for Zambia, based on the most recent Demographics and Health Survey for Zambia in 2018. This shows significant polarization to a younger population age structure. We note that the age distributions for the sampled and total population are very similar, supporting that our sample was representative of the larger population. By comparison, the CV19+ deaths show a relative increase in the deaths among older individuals and a relative decrease in younger age groups. While this tendency for deaths to be concentrated in the elderly is not unique, the extent of the skew towards older individuals is less pronounced from that seen in the US, the EU and China. In those populations, the proportion of deaths is virtually nil in those under age 50, and almost entirely concentrated in those 65 and above, with pediatric deaths being almost undetectably few. By contrast, 10% of the Lusaka deaths were in children.

**Figure 5a** overlays the CV19+ deaths by city ward, with higher numbers of cases indicated by darker coloring. The greatest numbers of cases were reported in the Kanyama, Chawama, Emmasdale and George wards. These wards are the most densely populated (**Figure 5b)** and least affluent areas of Lusaka.

**Figure 5.**
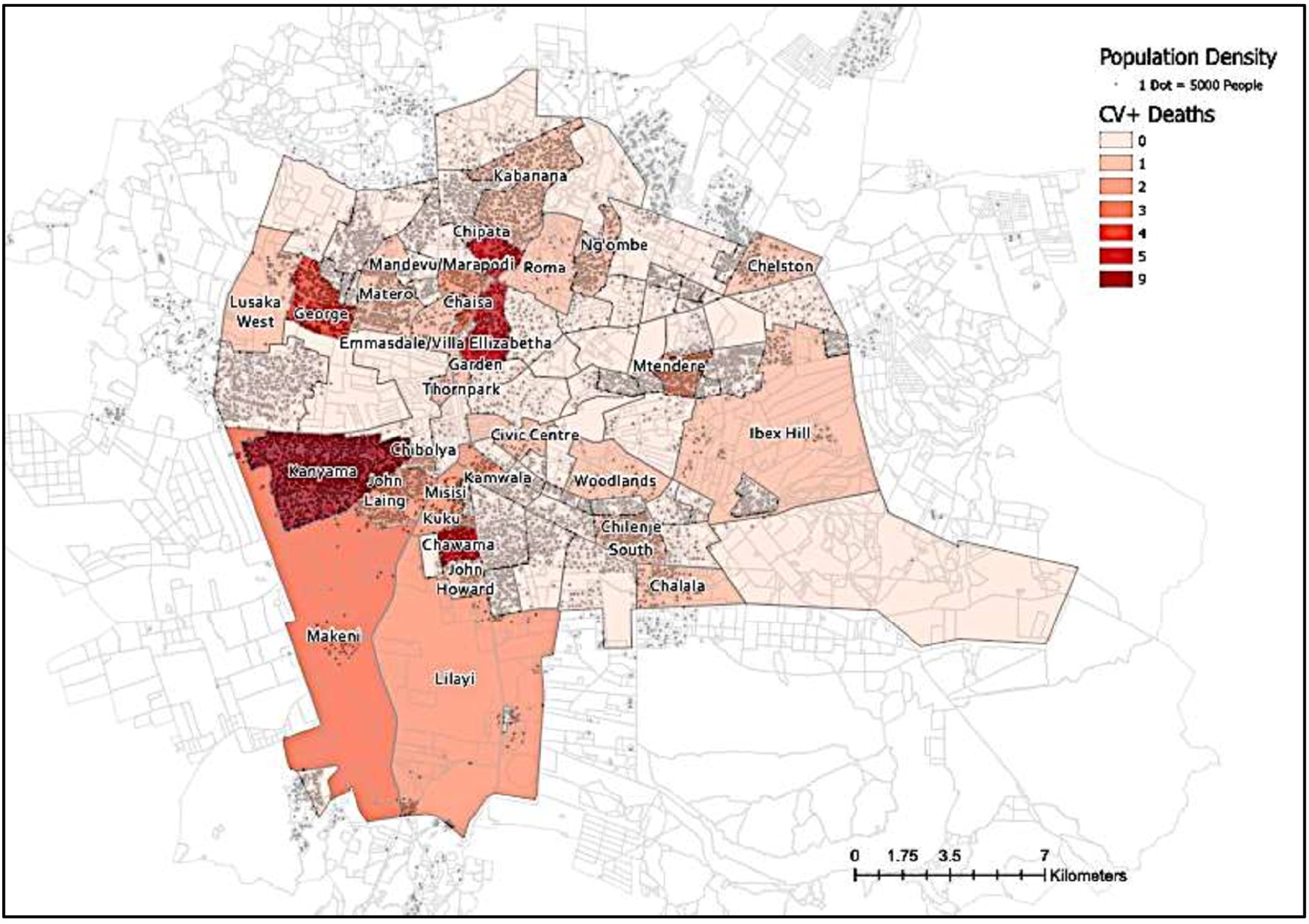
Geographic distribution of CV19 deaths by ward in Lusaka, Zambia. **Figure 5a.** Distribution of CV19 deaths by city ward We used the software ArcGIS to create this figure, importing data on population by city ward from numbers available on the Zambia data hub, which is managed by the Government of Zambia (see: https://zambia-open-data-nsdi-mlnr.hub.arcgis.com/). Each pip represents 5000 individuals. The heat map corresponds to the number of CV19 positive cases by referral clinic for the 70 CV19 positive deaths in our study. The four wards with the highest case burdens were the George, Chawama, Kanyama and Emmasdale compounds. These are also four of the poorest areas of Lusaka with the highest population and population densities.

**Figure 5b.**
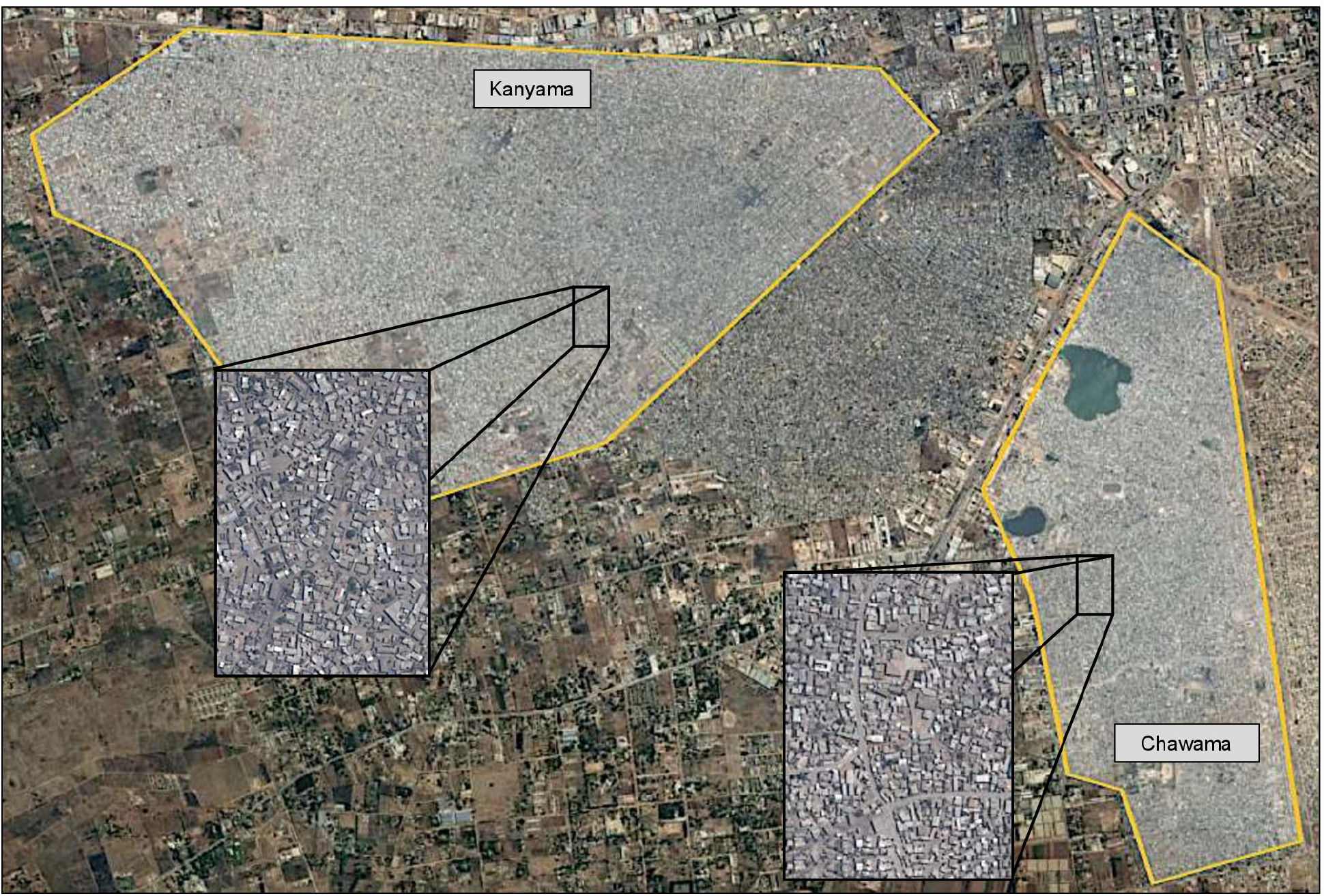
Satellite view of Lusaka highlighting the Kanyama and Chawama wards. These images were taken using Google Earth, highlighting the Kanyama and Chawama wards. These are the two largest population centers in Lusaka and the most impoverished. Even at lower magnification, the increased population density in these wards is readily apparent, as are the sharp delineations between adjacent more affluent areas of the city. The congestion becomes more apparent in the street-level inset figures.

### Clinical presentation of CV19 deaths

Most of the deceased presented with symptoms consistent with CV19 disease. We categorized 44/70 (66.7%) deaths as ‘probable CV19’ based on the presence of fever and/or respiratory symptoms, of which 16/44 (36%) were facility deaths and 28/44 (64%) were community deaths. Among the 19 facility deaths (for whom PCR testing ante-mortem could have been done), symptoms suggestive of CV19 were present among 17/19 (89.5%). The group of ‘probable CV19’ deaths included all six of the patients (5 adults and one child) who had been tested for CV19 antemortem.

A further 7/70 (10.0%), were considered ‘possible CV19’ given symptoms of sudden onset hemiparesis with or without preceding severe headaches, sudden onset left sided chest pain, or sudden onset abdominal pain preceding death. All but one of these were community deaths. The single facility death presented with headache on a background of anemia and chronic kidney disease.

A single case was deemed ‘probably not CV19’. This was an individual transferred for management of burns, where there was no mention of CV19 compatible symptoms.

This left 18/70 (25.7%) where causality remained ‘uncertain’ due to insufficient clinical data. All of these were community deaths. Thus for the 52 cases where data were sufficient to adjudicate causality, nearly all (51/52, 98.1%) presented with symptoms suggestive of, or compatible with, the CV19 syndrome.

### CV19 Deaths in children

While CV19 deaths in children were less common than other age groups, using the stricter CT threshold <40, we documented CV19 among five deaths in children <19 years. Two more children had detectable PCR signals with a CT ≥40 to <45, for a presumptive total of seven children who died with CV19. Of these, five were aged 3 years or less including three under the age of one year. The other two were teenagers. Only one of the children had been tested for CV19 antemortem (the 16 year old with epilepsy and developmental delay), with a negative PCR result at hospital admission and a positive result in our lab post-mortem. Clinical information on these individuals are provided in **Table 2**. In contrast with the adult population, who mainly presented with respiratory symptoms, among the children, gastrointestinal symptoms (nausea, vomiting, diarrhea or abdominal pains) were most common, and only one had respiratory symptoms. While the pediatric deaths account for only 10% of all CV19 deaths detected during this time, this ratio is nonetheless uncharacteristically high.

**Table 1.**
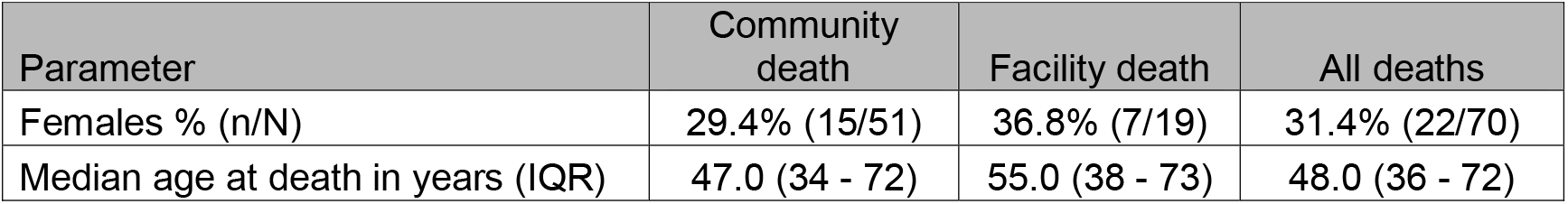
Demographic characteristics of CV19+ deaths in Lusaka, Zambia.

**Table 2.** Summary of pediatric deaths where CV19 was detected.

| Number | Clinical information | Location | Age range |
| --- | --- | --- | --- |
| **Detected at a PCR Cycle threshold <40** |  |  |  |
| 1 | Vomiting and diarrhea for two weeks. | Community | < 1 year |
| 2 | Sudden death – infant was reportedly well but found dead in the morning. | Community | < 1 year |
| 3 | Fever, difficulty breathing, nausea. | Facility | 1-5 years |
| 4 | Child collapsed suddenly while performing daily chores. No antecedent symptoms reported. | Community | 10-14 years |
| 5 | No reported acute symptoms, but the child had severe developmental delay and epilepsy and was non-verbal. | Community | 15-19 years |
| **Detected at a PCR cycle threshold 40 to <45** |  |  |  |
| 6 | Sore throat, headache, vomiting for six days* | Community | <1 years |
| 7 | Abdominal pains | Community | 1-5 years |
| * The young age of this child makes us skeptical of the history of 'sore throat and headache'. This can only be the assumption of the caregiver. By contrast, the vomiting episodes are easily observed and therefore more credible. |  |  |  |

### Prevalence of underlying risk factors among the CV19+ deaths

One or more potential underlying risk factors were identified in nearly all of the CV19 associated deaths in adults (**Table 3**). The most common conditions occurring in at least 10% of the cohort were, in declining order of prevalence: tuberculosis (31.4%), hypertension (27.1%), HIV/AIDS (22.9%), alcohol use (17.1%), and diabetes (12.9%). A recent diagnosis of malaria was mentioned several times. However, absent laboratory confirmation of this diagnosis, we interpreted this skeptically: malaria has effectively been eliminated from Lusaka yet still is often diagnosed on the basis of unexplained fever or malaise without laboratory confirmation, symptoms that could easily have been due to CV19 itself. Other than one child with epillepsy and developmental delay, none of the children had identified co-morbidities.

**Table 3.**
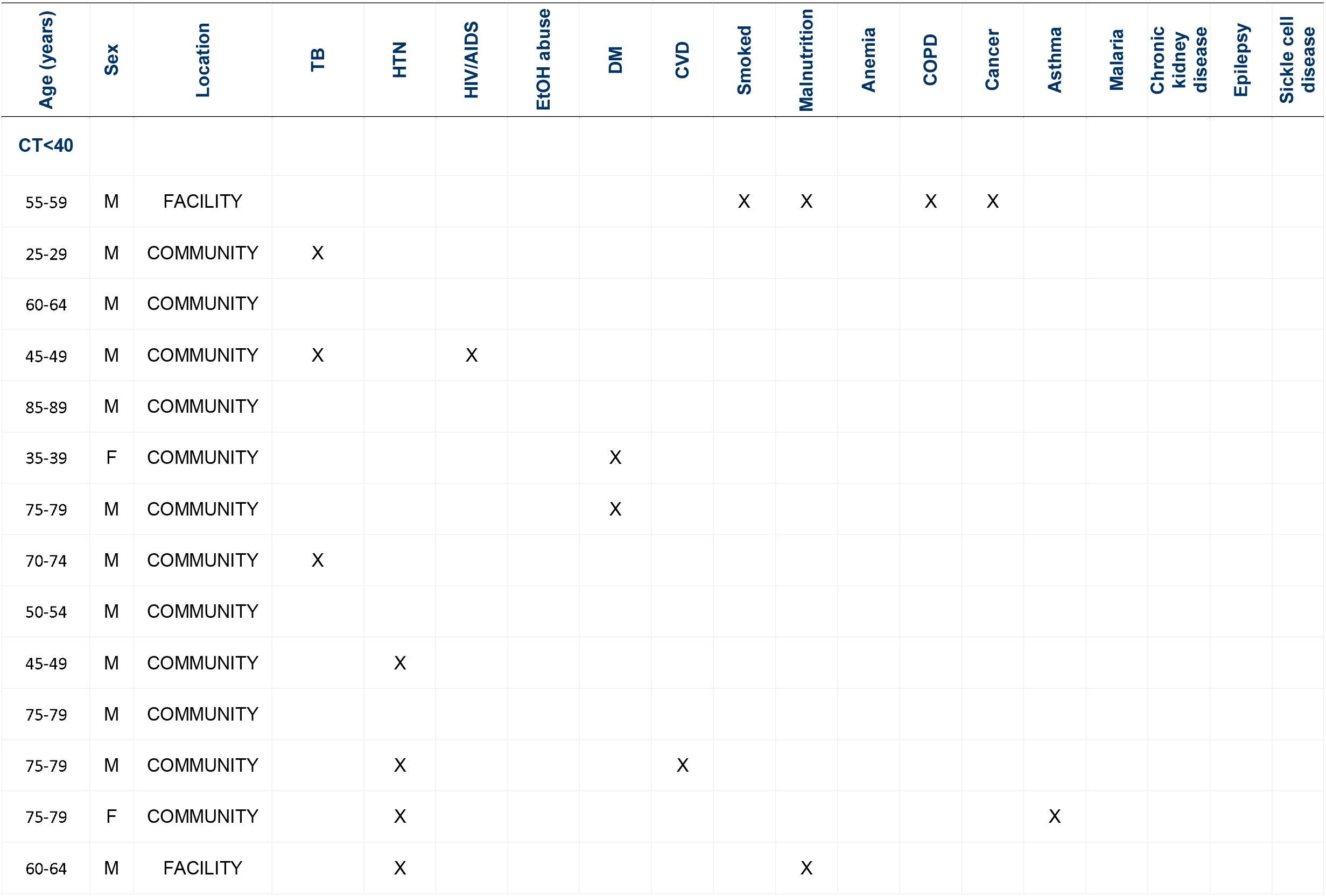
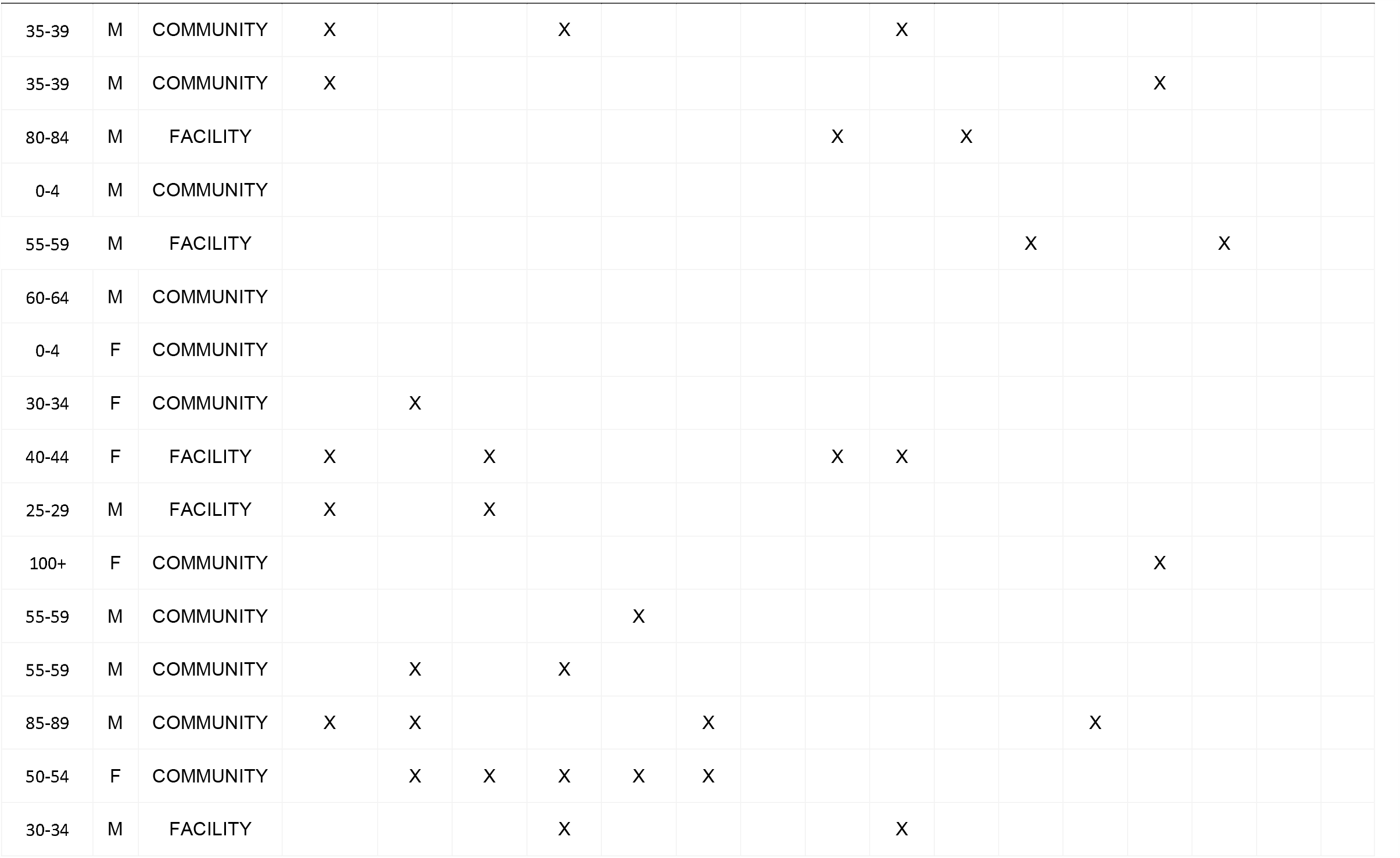
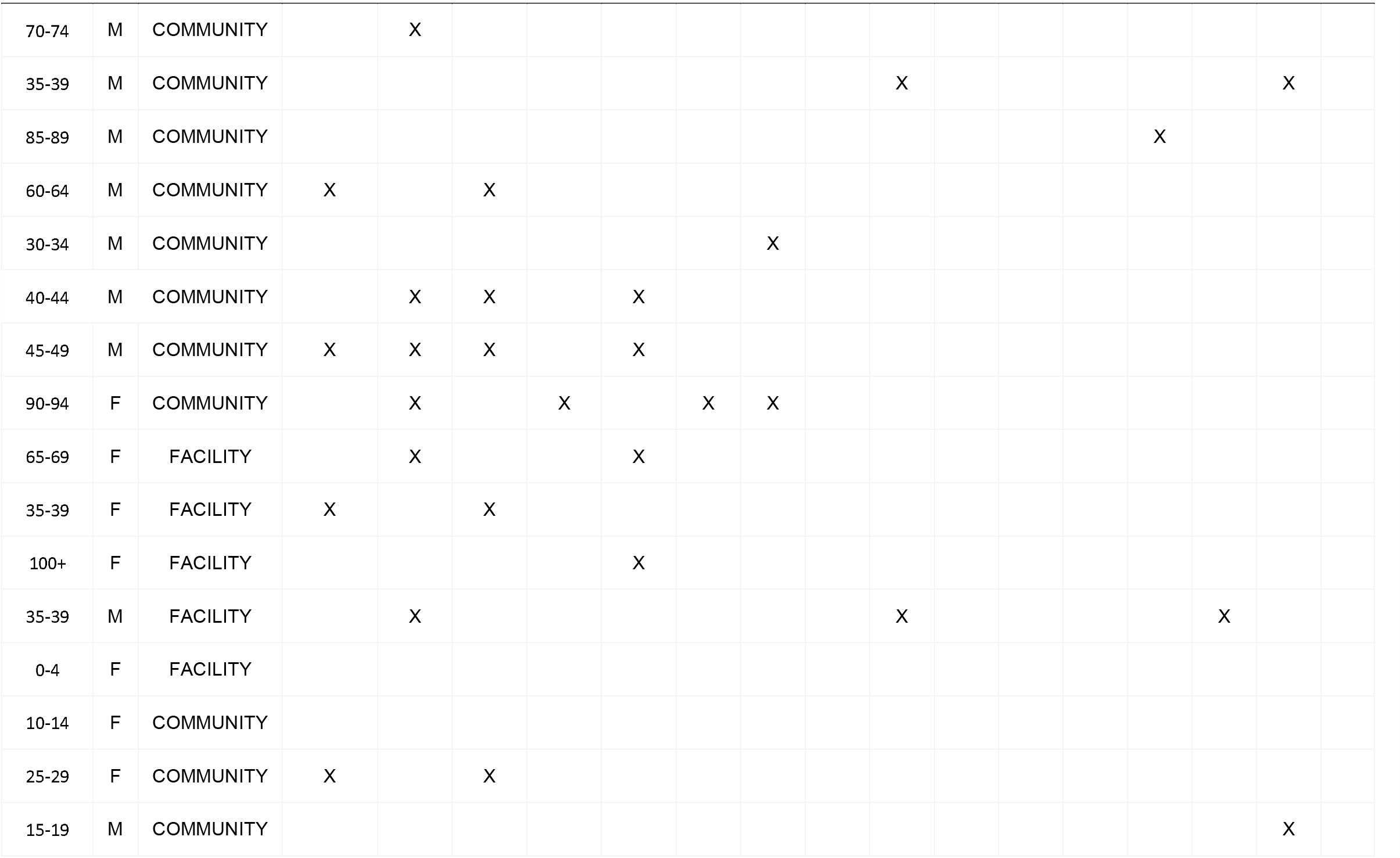
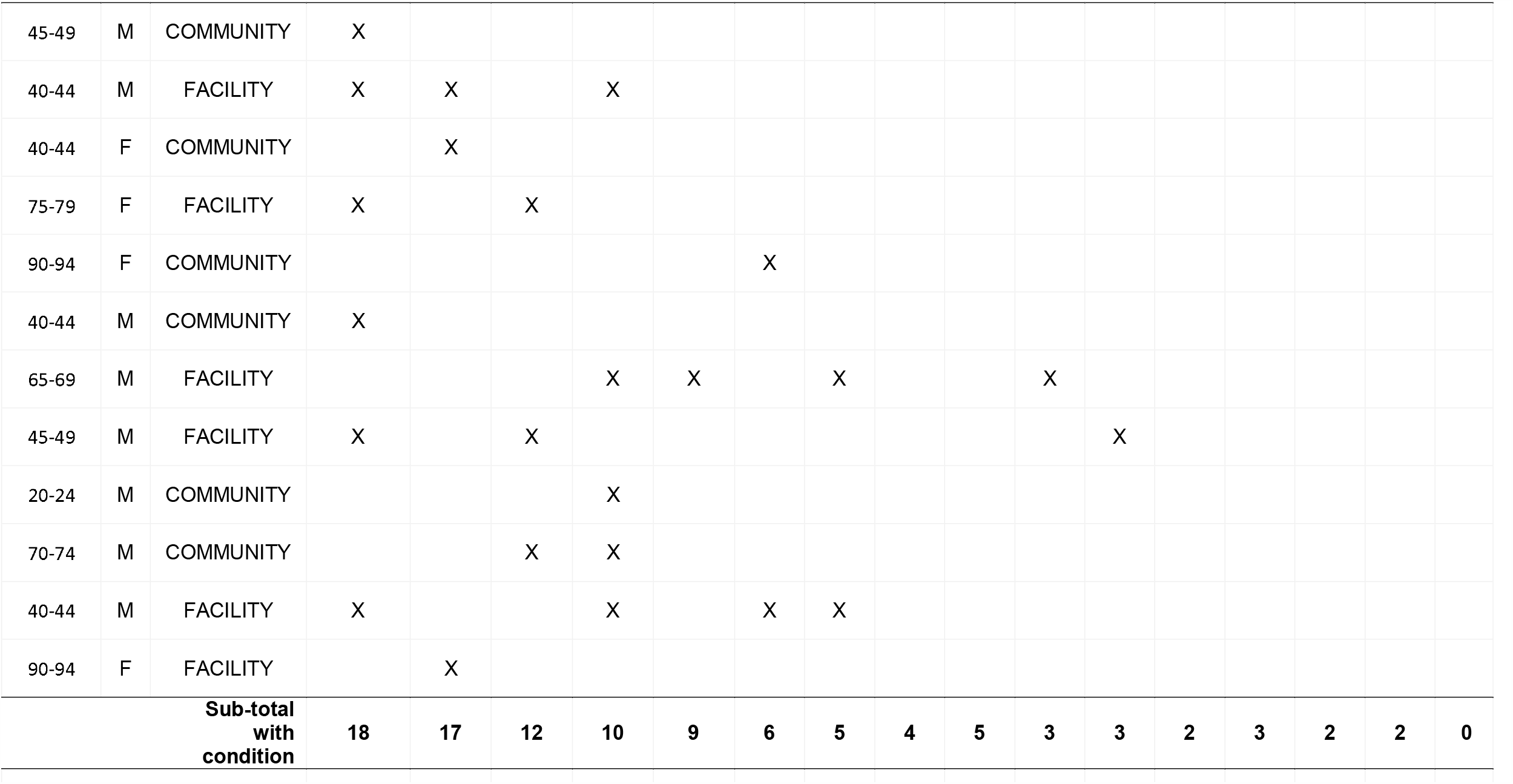
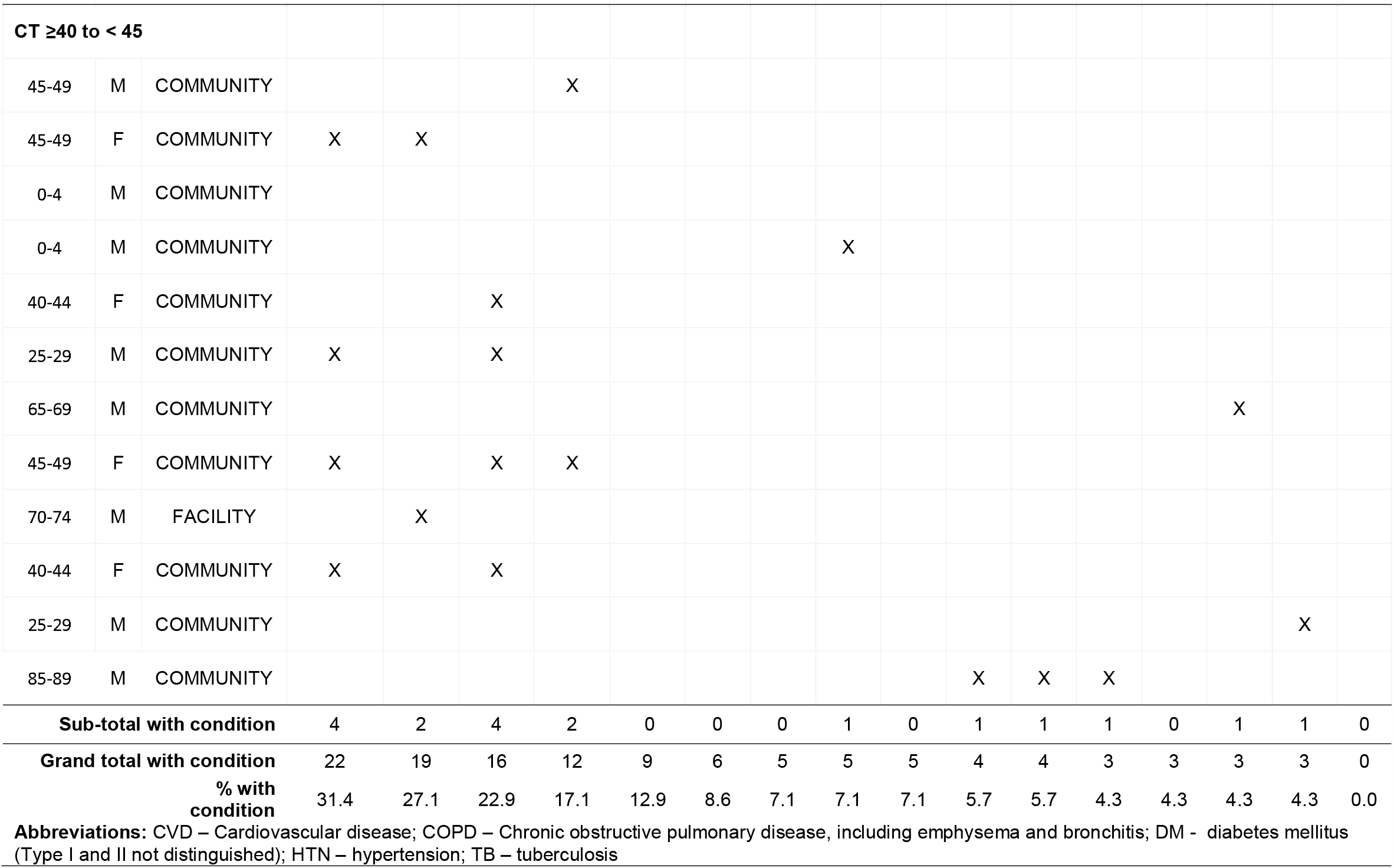
Prevalence of putative underlying risk factors for severe CV19 disease

## DISCUSSION

From this systematic surveillance study in Lusaka, Zambia, we observed a surprisingly high prevalence of CV19 mortality. Conservatively, 15% of all deceased individuals had CV19 during this period. If we assume that lower intensity PCR results (i.e., CT ≥40 to <45) reflect waning viral loads and are not false positives, then the CV19 prevalence approached 20% of all deaths. Only a minority had been tested for CV19 antemortem.

### Several points merit further discussion

First, most deaths occurred in the community and outside of medical care. Among the majority that were community deaths, none had been tested for CV19 antemortem. This is undoubtedly a significant factor in underestimating the impact of CV19 in Lusaka.

Second, testing was rarely conducted among the minority that were facility deaths, though nearly all had presented with a constellation of symptoms typical of CV19. We conclude that testing was neither widespread nor systematic.

Third, CV19 deaths occurred quite evenly across the age spectrum, not just among the elderly. In fact, most deaths were among indivduals aged 20-59 years. This pattern is distinct from that described in the US, the EU and China,^28-30^ and more typical of the death by age distributions and population age structures common in African countries. While there was a relative under-representation of CV19 in the younger population and a relative over representation in the older population, overall, the distribution of CV19+ deaths was similar to that seen in the burial registry cohort, therefore supporting the theory that the presentation of CV19 by age in this cohort is at least partially due to the expected distribution of deaths in Lusaka.

Fourth, ten percent (7/70) of the CV19+ deaths were in children, including three infants. Among the younger children, gastrointestinal rather than respiratory complaints predominated, which may be a factor explaining why only one had been tested for CV19 antemortem. In its November 19, 2020 update, the American Academy of Pediatrics reported that children accounted for between 0.00%-0.23% of all COVID-19 deaths in the United States, with 17 states reporting no child deaths.^31^ Given the extreme rarity of pediatric CV19 deaths in high income countries, we were surprised to observe any pediatric deaths in a group of seventy, let alone seven CV19+ deaths. This suggests another distinct feature of CV19’s presentation in Africa.

Fifth, we identified a high frequency of novel potential underlying risk factors that may be more specific to Africa. Notably, tuberculosis, malnutrition, and HIV/AIDS were very common in this setting but are comparatively uncommon in the US or Europe. These conditions are also highly correlated with each other. A possible association between CV19 and HIV/AIDS has been reported previously.^32 33^ While hypertension was the second most common risk factor, other chronic condictions such as diabetes, cardiovascular disease and emphysema were comparatively less frequent in the Lusaka cohort. These conditions are all common among older populations, and their relative infrequency here could be explained by Zambia’s younger population structure.

If our data are generalizable to other settings in Africa, the answer to the question, ‘*Why did CV19 skip Africa*?’ is that it didn’t.

To date, most of the systematically collected data about CV19 in Africa comes from South Africa, and those data suggest a substantial impact of CV19.^6-8^ But South Africa is a comparatively wealthy nation with significant resources and expertise in surveillance. This makes it unrepresentative of most other African nations, including Zambia. Elsewhere it is difficult to find evidence of widespread systematic testing in Africa. From sampling of blood donors, Kenya has reported seroprevalence rates among blood donors between 5-9%, even as PCR positivity was reportedly low.^34^ However, blood donors are probably not representative of the general population. As of November 24, 2020, the Nigerian CDC reported that only 0.3% of its population had been tested for CV19.^35^ Neither Nigeria nor Kenya reported the impact of CV19 in terms of hospitalizations or deaths. Across the continent, as of October 30, 2020, the Africa Centers for Disease Control reported a cumulative total of 21 million CV19 tests of which ∼60% came from only five of Africa’s 54 countries: South Africa, Morocco, Egypt, Kenya and Ethiopia.^36^ By contrast, the US, a country with less than one third the population of Africa, has conducted over 192 million CV19 tests, for a per capita rate nearly 30 times higher.^37^ Given these realities, an ‘absence of evidence’ documenting CV19’s impact in Africa could easily be misconstrued as ‘evidence of absence’.

Yet, understanding the true extent of CV19’s impact on Africa is critical. There is first a moral imperative that the world acknowledge suffering wherever it exists. But there are pragmatic reasons too. For one, if Africa’s citizens themselves view CV19 as posing little threat, they may less vigilant about taking actions that mitigate their personal risk, such as socially distancing or wearing masks. Likewise, this error accepted at a global level could put Africa at a lower priority for access to the forthcoming CV19 vaccines. The consequences of the misconception in both cases would be measured in lives.

Our analysis had several limitations. First, our data were collected over a short span of several months. Within this period, we saw dynamic shifts in the bi-weekly prevalence of CV19+ deaths. There is no way to predict how CV19’s patterns of transmission might shift in the future, which justifies our ongoing surveillance work. Second, our study could only infer links between deaths and CV19 when a PCR signal was detectable. This would fail to identify deaths indirectly due to CV19, such as heart attacks or strokes, that were separated in time from the CV19 infection. In an attempt to quantify the degree of excess mortality indirectly attributable to CV19, we are currently collecting age- and season-specific burial records from Lusaka from the past several years. Third, our assessment of underlying risk factors was limited by the completeness and accuracy of medical chart data and the possibly faulty recall of the next of kin. Hence, there was no way to verify these conditions, nor to assess their duration and severity. An analysis of the association between potential risk factors and CV19 deaths is currently underway in our group. Fourth, because the consequences of CV19 infection are age-specific, our data cannot be used to infer the incidence of CV19 in the larger population, but can only tell us about the prevalence of CV19 deaths across age groups. Lastly, our results came from one city, in one African country, over a short three-month span.

In conclusion, contradicting the prevailing narrative that CV19 has spared Africa, CV19 has had a severe impact in Zambia. How this was missed is largely explained by low testing rates, not by a low prevalence of CV19. Yet to avoid casting blame on Zambia, we note that the wealthiest countries on earth have struggled to test for and contain CV19. Can we be surprised that a resource poor country would also struggle to respond effectively to the greatest global public health emergency since HIV/AIDS? Establishing systematic disease surveillance requires time and significant resources. Zambia had neither. The challenge of scarce resources is hardly unique to Zambia, and Zambia is hardly the poorest nation in Africa. If our data are generalizable, CV19’s impact across Africa has been substantially underestimated.

## Data Availability

We are willing to share our data

## ACKNOWLEDGEMENTS

We wish to dedicate this paper to the memory of our beloved team member Dr. Roy Chavuma. Roy was one of countless many who were lost to CV19 this year. He was an outstanding surgeon, a devoted public heatlh professional, and an inspired mentor to dozens of young clinicians and future scientists over many decades of service. But he was primarily our dear friend. His laughter and optimism kept our spirits high in the darkest of times. He will be missed but not forgotten.

The ZPRIME study and the CV19 expansion were made possible through the generous support of the Bill & Melinda Gates Foundation (OPP 1163027).

## Notes

### Competing Interest Statement

The authors have declared no competing interest.

### Clinical Trial

This was not a clinical trial

### Funding Statement

The work was funded by the Bill & Melinda Gates Foundation.

### Author Declarations

This research protocol was approved by the ethical review boards at Boston University Medical Center and The University of Zambia.

## REFERENCES

1. Viglione G. How many people has the coronavirus killed? Nature 2020;585(7823):22–24. doi: 10.1038/d41586-020-02497-w [published Online First: 2020/09/03]

2. Bamgboye EL, Omiye JA, Afolaranmi OJ, et al. COVID-19 Pandemic: Is Africa Different? J Natl Med Assoc 2020 doi: 10.1016/j.jnma.2020.10.001 [published Online First: 2020/11/07]

3. Lawal Y. Africa’s low COVID-19 mortality rate: A paradox? Int J Infect Dis 2020;102:118–22. doi: 10.1016/j.ijid.2020.10.038 [published Online First: 2020/10/20]

4. Twahirwa Rwema JO, Diouf D, Phaswana-Mafuya N, et al. COVID-19 Across Africa: Epidemiologic Heterogeneity and Necessity of Contextually Relevant Transmission Models and Intervention Strategies. Ann Intern Med 2020;173(9):752–53. doi: 10.7326/M20-2628 [published Online First: 2020/06/20]

5. Troeger C. Just how do deaths due to COVID-19 stack up? In: Health TG, ed., 2020.

6. Abdool Karim SS. The South African Response to the Pandemic. N Engl J Med 2020;382(24):e95. doi: 10.1056/NEJMc2014960 [published Online First: 2020/05/30]

7. Nordling L. ‘Our epidemic could exceed a million cases’ - South Africa’s top coronavirus adviser. Nature 2020;583(7818):672. doi: 10.1038/d41586-020-02216-5 [published Online First: 2020/07/29]

8. National Institutes for Communicable Diseases (NICD). COVID-19 weekly epidemiology brief. Johannesburg, S. Africa: National Institutes of Communicable Diseases, 2020.

9. Nordling L. Africa’s pandemic puzzle: why so few cases and deaths? Science 2020;369(6505):756–57. doi: 10.1126/science.369.6505.756 [published Online First: 2020/08/15]

10. Mbow M, Lell B, Jochems SP, et al. COVID-19 in Africa: Dampening the storm? Science 2020;369(6504):624–26. doi: 10.1126/science.abd3902 [published Online First: 2020/08/09]

11. Ntoumi F, Velavan TP. COVID-19 in Africa: between hope and reality. Lancet Infect Dis 2020 doi: 10.1016/S1473-3099(20)30465-5 [published Online First: 2020/06/20]

12. Lancet editorial staff. COVID-19 in Africa: no room for complacency. Lancet 2020;395(10238):1669. doi: 10.1016/S0140-6736(20)31237-X [published Online First: 2020/06/01]

13. El-Sadr WM, Justman J. Africa in the Path of Covid-19. N Engl J Med 2020;383(3):e11. doi: 10.1056/NEJMp2008193 [published Online First: 2020/04/18]

14. Shrock E, Fujimura E, Kula T, et al. Viral epitope profiling of COVID-19 patients reveals cross-reactivity and correlates of severity. Science 2020 doi: 10.1126/science.abd4250 [published Online First: 2020/10/01]

15. Bell D, Hansen KS, Kiragga AN, et al. Predicting the Impact of COVID-19 and the Potential Impact of the Public Health Response on Disease Burden in Uganda. Am J Trop Med Hyg 2020;103(3):1191–97. doi: 10.4269/ajtmh.20-0546 [published Online First: 2020/07/25]

16. Ditekemena J. COVID-19 amidst Ebola’s retreat. Science 2020;368(6490):445. doi: 10.1126/science.abc4859 [published Online First: 2020/05/02]

17. Thompson KM, Kalkowska DA, Badizadegan K. A Health Economic Analysis for Oral Poliovirus Vaccine to Prevent COVID-19 in the United States. Risk Anal 2020 doi: 10.1111/risa.13614 [published Online First: 2020/10/22]

18. Ogimi C, Qu P, Boeckh M, et al. Association between live childhood vaccines and COVID-19 outcomes: a national-level analysis. medRxiv 2020 doi: 10.1101/2020.10.17.20214510 [published Online First: 2020/10/28]

19. Ozdemir O. Measles-Mumps-Rubella Vaccine and COVID-19 Relationship. Mbio 2020;11(5) doi: 10.1128/mBio.01832-20 [published Online First: 2020/09/24]

20. The World Bank. The World Bank in Zambia - Overview 2020 [Available from: https://www.worldbank.org/en/country/zambia/overview.

21. The Central Intelligence Agency. The World Fact Book - Zambia, 2020.

22. Zambia Statistic’s Agency (formerly the Central Statistical Office). Zambia Demographic and Health Survey 2018, 2018.

23. Serina P, Riley I, Stewart A, et al. A shortened verbal autopsy instrument for use in routine mortality surveillance systems. BMC Med 2015;13:302. doi: 10.1186/s12916-015-0528-8

24. Van Horn KG, Audette CD, Tucker KA, et al. Comparison of 3 swab transport systems for direct release and recovery of aerobic and anaerobic bacteria. Diagn Microbiol Infect Dis 2008;62(4):471–3. doi: 10.1016/j.diagmicrobio.2008.08.004 [published Online First: 2008/09/26]

25. Dube FS, Kaba M, Whittaker E, et al. Detection of Streptococcus pneumoniae from Different Types of Nasopharyngeal Swabs in Children. PLoS One 2013;8(6):e68097. doi: 10.1371/journal.pone.0068097 [published Online First: 2013/07/11]

26. Yang G, Erdman DE, Kodani M, et al. Comparison of commercial systems for extraction of nucleic acids from DNA/RNA respiratory pathogens. J Virol Methods 2011;171(1):195–9. doi: 10.1016/j.jviromet.2010.10.024 [published Online First: 2010/11/03]

27. Centers for Disease Control and Prevention. Guidance for certifying deaths due to coronavirus disease 2919 (COVID-19). Vital Statistics Reporting Guidance. Hyattsville, Maryland: National Center for Health Statistics, 2020.

28. Zhang J, Litvinova M, Wang W, et al. Evolving epidemiology and transmission dynamics of coronavirus disease 2019 outside Hubei province, China: a descriptive and modelling study. Lancet Infect Dis 2020;20(7):793–802. doi: 10.1016/S1473-3099(20)30230-9 [published Online First: 2020/04/06]

29. Giacomelli A, Ridolfo AL, Milazzo L, et al. 30-day mortality in patients hospitalized with COVID-19 during the first wave of the Italian epidemic: A prospective cohort study. Pharmacol Res 2020;158:104931. doi: 10.1016/j.phrs.2020.104931 [published Online First: 2020/05/25]

30. Bhatraju PK, Ghassemieh BJ, Nichols M, et al. Covid-19 in Critically Ill Patients in the Seattle Region - Case Series. N Engl J Med 2020;382(21):2012–22. doi: 10.1056/NEJMoa2004500 [published Online First: 2020/04/01]

31. American Academy of Pediatrics (AAP). Children and COVID-19: State Data Report: American Academy of Pediatrics, November 16, 2020.

32. Akyala AI, Iwu CJ. Novel severe acute respiratory syndrome coronavirus 2 (SARS-CoV-2) co-infection with HIV: clinical case series analysis in North Central Nigeria. Pan Afr Med J 2020;37:47. doi: 10.11604/pamj.2020.37.47.24200 [published Online First: 2020/11/20]

33. Parker A, Koegelenberg CFN, Moolla MS, et al. High HIV prevalence in an early cohort of hospital admissions with COVID-19 in Cape Town, South Africa. S Afr Med J 2020;110(10):982–87. doi: 10.7196/SAMJ.2020.v110i10.15067 [published Online First: 2020/11/19]

34. Ojal J BS, Were V, Okiro EA, Kmobe IK, Mburur C, Aziza R, Ogero M, Agweyu A. Marimwe GM, Uyoga S, Adteifa IMO, Scott JA, Otieno E, Ochola-Ohier LI, Agoti CN, Kasera K, Amoth P, Mwangangi M, Aman R, Ng’ang’a W, Tsofa B, Bejon P, Barasa E, Keeling MJ, Nokes DJ. Revealing the extent of teh COVID-19 pandemic in Kenya based on serological and PCR-test data. medRxiv 2020 [published Online First: September 03, 2020]

35. Nigerian Centers for Disease Control. COVID-19 - Nigeria November 2020 [cited 2020 24 November 2020]. Available from: https://covid19.ncdc.gov.ng/.

36. Africa Centers for Disease Control. Laboratory Technical Working Group The African Union, 2020.

37. The COVID Tracking Project. US Historical Data 2020 [30 November 2020]. Available from: https://covidtracking.com/data/national.

